# Changes to hypothalamic volume and associated subfields during gender-affirming hormone treatment in gender dysphoria

**DOI:** 10.1101/2022.02.02.22270319

**Authors:** ME Konadu, MB Reed, U Kaufmann, PA Handschuh, M Spies, B Spurny-Dworak, M Klöbl, V Ritter, GM Godbersen, R Seiger, P Baldinger-Melich, GS Kranz, R Lanzenberger

## Abstract

Sex steroid hormones influence hypothalamic micro- and macrostructure in humans and animal models. Neuroimaging studies have suggested that estrogen and anti-androgen treatment decreases volumes of multiple cortical and subcortical brain areas in transgender individuals, including total hypothalamus volume. Here, we aim to further explore potential effects of gender-affirming hormone treatment (GHT) in transgender individuals on hypothalamic volume by providing additional information on hypothalamic subfields.

38 transgender men (TM) and 15 transgender women (TW), with gender dysphoria (DSM-5), as well as 32 cisgender women (CW) and 21 cisgender men (CM) underwent two magnetic resonance imaging (MRI) measurements with an interval of at least four months (median interval TM= 134.5 days (interquartile range (IQR): 126-152.25); TW= 149 days (IQR: 126-178.5); CW= 147 days (IQR: 139.75-170.5); CM= 146 days (IQR: 132-247)) between both sessions. In transgender individuals GHT, consisting of estrogen and anti-androgen treatment in TW and testosterone treatment in TM, was initiated directly after the first measurement. To assess how GHT interacts with hypothalamic structures, the hypothalamus and its subunits were segmented using FreeSurfer. Subject group x time interaction effects were evaluated using repeated measures ANCOVA models. The Bonferroni method was used to correct for multiple comparisons.

Significant decreases of total hypothalamic volume and associated subunits were detected in TW after estrogen and anti-androgen treatment compared to cisgender groups. Effects were found in the total hypothalamus volume (p_corr_= 0.001), the left and right hypothalamus (p_corr_= 0.002), the inferior tubular subunit bilaterally (right: p_corr_= 0.001; left: p_corr_= 0.001), the left superior tubular subunit (p_corr_= 0.003) the right anterior inferior subunit (p_corr_= 0.002), as well as the right anterior superior subunit (p_corr_= 0.0002) of the hypothalamus.

Here, we observed significant volumetric effects on the adult human hypothalamus after an interval of at least four months of estrogen and anti-androgen treatment in TW and added knowledge on associated subfields. Further studies investigating influences of sex steroid hormones on brain structure and functional connections are still needed.

## 2. Introduction

The hypothalamus is a subcortical structure in the human brain that acts as a neuroendocrine control center of the human body. It is subdivided into several nuclei, each of which exert specific actions (Swaab et al., 1993), ranging from the control of body temperature (Zhao et al., 2017), and energy homeostasis (Timper & Brüning, 2017), to sexual (Paredes, 2003), and aggressive behavior (Gouveia et al., 2019). Sex steroid hormones are believed to influence brain development in the perinatal brain, as well as in adulthood by so-called “organizational” and “activational” effects (Arnold, 2009). Sex differences in the hypothalamus are thought to mainly be driven by the effects of estradiol (Lenz & McCarthy, 2010) and have been observed in rodent studies in several hypothalamic substructures including the arcuate nucleus, preoptic area and the medial basal hypothalamus (Brawer et al., 1983; Brawer et al., 1978; Lenz & McCarthy, 2010). A recent review emphasized that a substructure of the human hypothalamus, the third interstitial nucleus of the anterior hypothalamus (INAH-3), displays the largest difference of human brain volumes between cisgender male and female individuals (Eliot et al., 2021). This is supported by an earlier study, which revealed larger hypothalamus volumes in cisgender men and linked these findings to higher amounts of estrogen and androgen receptor densities in this structure (Goldstein et al., 2001). Interestingly, when analyzing central effects of oral contraceptives (OCP), mainly consisting of estrogens and progestogens (Dragoman, 2014), a reduction of hypothalamic grey matter volume in OCP users compared to naturally cycling women was revealed (Chen et al., 2021). Therefore, an influence of sex steroid hormones on hypothalamic structure could be assumed. Furthermore, it has been found that hypothalamic volumes show variations in affective disorders (Schindler et al., 2012; Schindler et al., 2019). Mood disorders are known to have different prevalence rates in males and females, which in part might be influenced by hormones (Salk et al., 2017). Considering these aspects, the hypothalamus represents a highly relevant target when investigating the impact of sex steroid hormones on the human living brain.

A unique approach to study direct effects of high dosages of sex steroid hormones on the human brain is to investigate transgender individuals undergoing gender-affirming hormone treatment (GHT) (Kranz et al., 2020). The term “gender dysphoria” (DSM-5), or “gender incongruence” (ICD-11) describes an individual’s feeling of incongruence between the experienced gender identification, “gender identity” and the biological gender assigned at birth. GHT typically consists of estrogen and anti-androgen treatment in transgender women (TW; male assigned at birth, with female gender identity) and testosterone treatment in transgender men (TM; female assigned at birth, with male gender identity) (Kranz et al., 2020) and is linked to improved mental health outcomes and reductions in body dissatisfaction (Nguyen et al., 2019).

A previous study by our group investigating the impact of GHT on hypothalamic microstructure assessed by diffusion-weighted imaging (DWI) found androgen-related reductions of the mean diffusivity (MD) in the lateral hypothalamus of TM receiving testosterone replacement therapy for four months (Kranz et al., 2018). These findings suggest a change of hypothalamic microstructure towards proportions found in cisgender men (CM; male assigned at birth, with male gender identity) (Kranz et al., 2018). Another in vivo neuroimaging study investigated structural changes of grey matter volumes before and after four months of GHT in a sample of eight TM and six transgender women. In TM, androgen treatment led to an increase of total brain volume into the direction observed in CM. Anti-androgen and estrogen treatment in TW, on the other hand, led to a decline of total brain and hypothalamus volumes towards values typically found in cisgender women (CW; female assigned at birth, with female gender identity) (Pol et al., 2006). Also, in post-mortem studies of transgender persons who have undergone GHT in their lives, the INAH-3 in TW has been found to be comparable to that of CW in terms of volume and number of neurons. In one TM subject the INAH-3 size was comparable to sizes of CM (Garcia-Falgueras & Swaab, 2008).

Neuroimaging studies investigating the effects of GHT on the brain, specifically on the hypothalamus, are scarce and mostly underpowered. Our study aims to further elucidate the potential link between sex steroid hormones and morphologic changes of the adult human brain. Therefore, we examined hypothalamic volume, including its subfields before and after four months of GHT in transgender individuals. Based on existing findings, we hypothesized a decrease of total hypothalamic volume in TW after four months of GHT.

## 3. Methods

### 3.1. Sample

The data analyzed was combined from two larger studies conducted by our group and partly has been published earlier addressing other research questions (Baldinger-Melich et al., 2020; Hahn et al., 2016; Kranz et al., 2014; Kranz et al., 2018; Kranz et al., 2017; Seiger et al., 2016; Spies et al., 2016). For this analysis, we considered data from 125 right-handed subjects who underwent two magnetic resonance imaging (MRI) measurements in a longitudinal design. 19 subjects had to be excluded before analysis because of poor MRI data quality. The resulting sample of 106 participants included into the final analysis, consisted of 32 CW (mean age ± standard deviation (SD)= 25.14 ± 6.06), 38 TM (mean age ± SD= 25.94 ± 6.49), 21 CM (mean age ± SD= 26.62± 6.41) and 15 TW (mean age ± SD= 26.26 ± 4.26) participants. Two MRI measurements were performed in all subjects with a median interval of 134.5 days in TM (interquartile range (IQR): 126-152.25), 149 days in TW (IQR: 126-178.5), 147 days in CW (IQR: 139.75-170.5) and 146 days in CM (IQR: 132-247). GHT was initiated in transgender individuals directly after the first measurement.

Transgender subjects were recruited from the transgender outpatient unit of the Department of Obstetrics and Gynecology, Unit for Gender Identity Disorder, Medical University of Vienna. A diagnosis of gender dysphoria according to the Diagnostic and Statistical Manual of Mental Disorders (DSM)-5, designated as gender incongruence in the International Classification of Diseases (ICD)-11, and the desire for GHT had to be present for enrollment. Cisgender subjects were recruited via online media as well as flyers on designated message boards in Vienna and the Medical University of Vienna. Participants with presence or history of a physical, neurological, or psychiatric disorder (cisgender subjects) or a major DSM-5 Axis-I comorbidity (transgender subjects), abnormal blood values or anomalies in clinical examinations were excluded from the study. Additionally, pregnancy, breastfeeding, substance abuse (except smoking), steroid hormone treatment within six months prior to study start, and contraindications for the participation in MRI measurements were set as exclusion criteria.

Both studies were approved by the Ethics Committee of the Medical University of Vienna (EK number study 1: 1104/2015, study: 644/2010) and registered at ClinicalTrials.gov (ClinicalTrials.gov Identifier study 1: NCT02715232; study 2: NCT01292785). Every subject gave written informed consent, was insured and received financial reimbursement for participation. All procedures performed during both studies were in accordance with the Helsinki declaration and latest revisions.

### 3.2. Study Design and treatment protocol

Both studies were conducted as longitudinal mono-center studies. All participants underwent a baseline MRI scan and a second MRI scan after an interval of at least four months (median intervals see section 3.1). Directly after the first MRI measurement GHT was initiated in transgender subjects. Checkups of the hormone treatment were performed regularly for all transgender participants at the Department of Obstetrics and Gynecology, Unit of Gender Identity Disorder of the Medical University of Vienna.

TM received either 1000 mg of testosterone undecanoate administration every 8-12 weeks (Nebido^®^, 250 mg/ml, 4 ml vial, applied intramuscularly) or 50 mg testosterone cream or gel applied transdermally (Testogel^®^, Testavan^®^). Additionally, in some cases 75 µg desogestrel (Moniq Gynial^®^) per day or triptorelin acetate as a depot (Decapeptyl^®^) was given every 4-5 weeks. TW received 25 to 50 mg cyproterone acetate daily (Androcur^®^) and estradiol 2×2 mg per day (Estrofem^®^) or estradiol 1.5-3 mg per day applied transdermally (Estrogel^®^) was administered. In some cases, triptorelin acetate as a depot (Decapeptyl^®^) was given every 4-5 weeks. Some participants additionally received progesterone (Arefam^®^) 100-200mg per day. If extensive hair loss occurred, 2.5 mg of an alpha-5-reductase-inhibitor (Finasterid) every second day was prescribed.

### 3.3. Hormonal blood sampling

Hormonal blood samples were drawn at each MRI measurement from all participants. The plasma levels of dehydroepiandrosterone sulfate (DHEAS), 17β-estradiol, follicle-stimulating hormone (FSH), luteinizing hormone (LH), progesterone, sex hormone-binding globulin (SHBG) and testosterone were analyzed by the Department of Laboratory Medicine, Medical University of Vienna (http://www.kimcl.at). These hormonal values have been included in our statistical analysis to correct for different hormonal treatment regimens transgender participants received, as well as for unknown effects of other relevant hormonal values.

### 3.4. Data acquisition

As the data analyzed was combined from two studies conducted by our group, MRI measurements were performed using two separate 3 Tesla MRI scanners (study 1: Siemens Magnetom Prisma, Erlangen, Germany/ study 2: Siemens Tim Trio, Erlangen, Germany) with a 64-channel (study 1) and 32-channel (study 2) head coil and a T1 weighted magnetization prepared rapid gradient echo (MPRAGE) sequence (study 1: TE= 2.91ms; TR=2000ms; TI= 900ms; *α*= 9°; 192 slices; 240 × 256 matrix; 1 × 1 × 1 mm^3^ voxel size/ study 2: TE= 4.21ms; TR= 2300ms; TI= 900ms, *α*= 9°; 160 slices; 240 × 256 matrix; 1.1 × 1 × 1 mm^3^ voxel size, total acquisition time 7 min, 46s). LongCombat was employed to correct for means and variances of the residuals across different scanners in this longitudinal setting (Beer et al., 2020). Using this method, the group and time differences were set to be maintained, while the scanner differences were harmonized.

### 3.5. Data processing

A visual quality check of the MRI data was performed. Subjects were processed with the standard pipeline of the FreeSurfer software suit (http://surfer.nmr.mgh.harvard.edu/, version 7.2) (Dale et al., 1999; Fischl et al., 1999). Our study model consists of measurements at two timepoints, therefore the longitudinal stream was used afterwards (Reuter et al., 2012). Subsequently, the hypothalamus and its subfields were segmented using the recently presented automated segmentation tool for the hypothalamus and associated subfields, which is distributed with FreeSurfer (Billot et al., 2020). In this tool hypothalamic subnuclei are assigned to five hypothalamic subunits on each side, which are classified according to *Bocchetta et al. and Makris et al*. (Bocchetta et al., 2015; Makris et al., 2013). The automated segmentation has been checked visually by a trained neuroscientist.

### 3.6. Statistical analysis

Statistical analysis was performed using SPSS version 25 for Windows (SPSS Inc., Chicago; Illinois; www.spss.com). Repeated measures analysis of covariance (rmANCOVA) was used to investigate the influence of GHT on hypothalamic volumes. Group (CW, TM, CM, TW) was used as between-subject-factor, whereas time (M1, M2) was included as the within-subject-factor. The interaction of group x time was examined in a stepwise approach. A separate analysis of the total hypothalamic volume was performed, if significant each side was analyzed, and if significant again, each subunit in the respective side was tested. To correct for different hormonal treatment regimens, as well as effects of other relevant hormonal values, we included DHEAS, SHBG, FSH, LH, progesterone, estradiol and testosterone of each participant into our analysis as covariates. To reduce the dimensionality of these values, a principal component analysis (PCA) was applied and the first two components explaining >98% of the total variance were integrated into our statistical model as covariates. The total intracranial volume (TIV) was also included as a covariate into our model (Eliot et al., 2021). The Bonferroni method was used to correct for multiple comparisons of regions and post-hoc tests included in each analysis.

## 4. Results

Analyses revealed an interaction effect of group x time in the total volume of the hypothalamus (F=6.281, p_corr_=0.001). When analyzing the left (F=6.397, p_corr_=0.002) and right (F=6.080, p_corr_=0.002) hypothalamus, again significant interaction effects of group x time were found. In the single nuclei, significant interaction effects of group x time were found in the left inferior tubular subunit (F=7.267, p_corr_=0.001), right inferior tubular subunit (F=7.294, p_corr_=0.001), left superior tubular subunit (F=6.689, p_corr_=0.003), right anterior superior subunit (F=7.025, p_corr_=0.0002) and right anterior inferior subunit (F=7.387, p_corr_=0.002). Post-hoc pairwise comparison between M1 and M2 in each group displayed a significant volume decrease in these regions in the TW group compared to cisgender groups (see Fig.1). All results reported were significant after correcting for multiple comparisons using the Bonferroni method for the number of regions and post-hoc tests included in each analysis. In CW smaller hypothalamus volumes have been observed at the second measurement compared to the first measurement, which, however, did not reach significance. Also, in TM and CM no significant differences between the two timepoints were detected. Significant results found in TW are summarized in Table 1.

**Figure 1:**
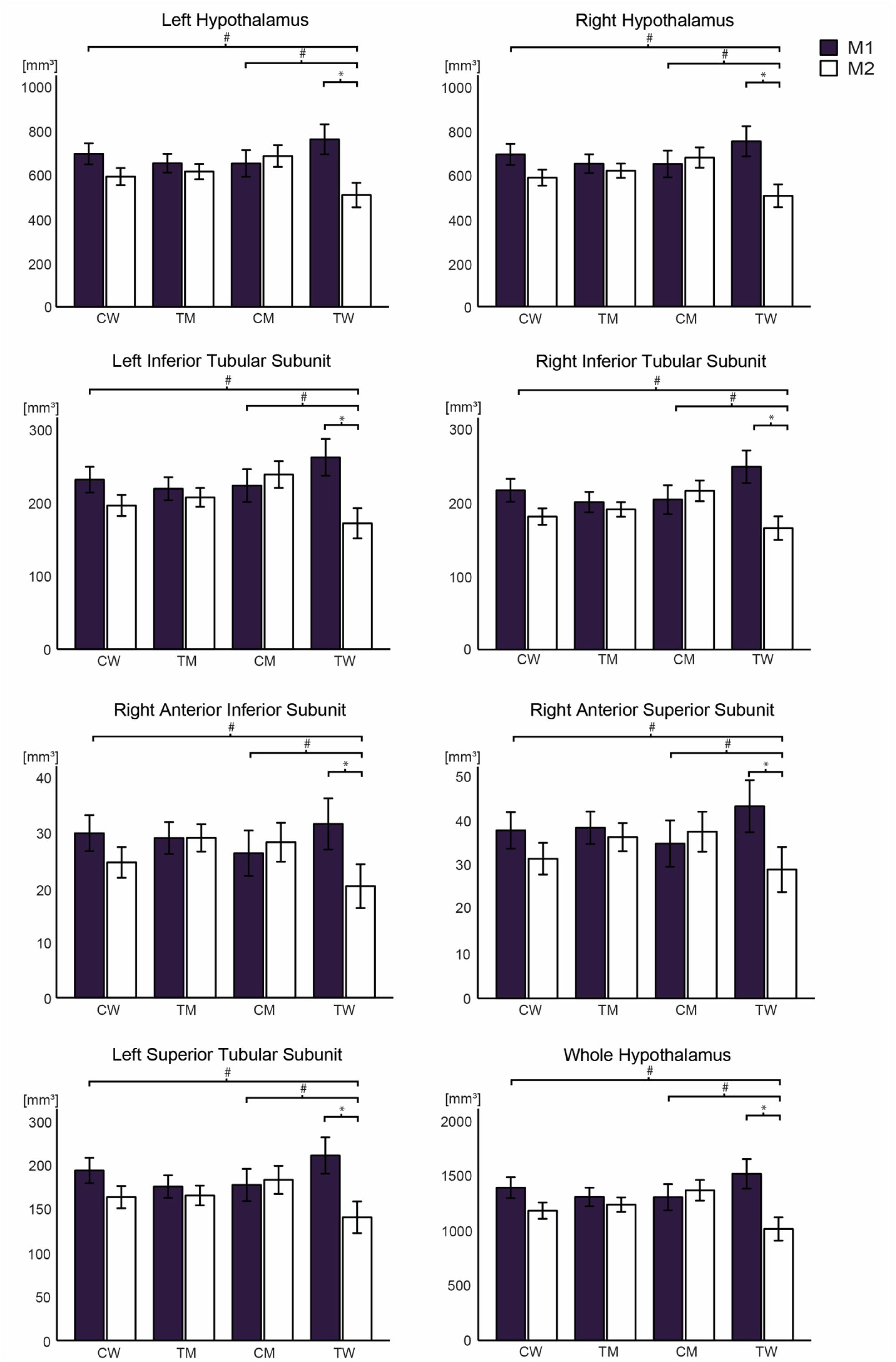
Decreased volumes of the hypothalamus and associated subfields in TW after an interval of at least four months of GHT. Post-hoc comparisons between baseline MRI (M1) and MRI after an interval of at least four months (M2) in each subject group (CW, TM, CM, TW). Transgender participants received GHT directly after the first measurement. A significant reduction of hypothalamic volumes as well as subfields is given in the TW group in comparison to cisgender groups (whole hypothalamus, left and right hypothalamus, left and right inferior tubular subunit, right anterior inferior subunit, right anterior superior subunit, left superior tubular subunit). (*) reports significant results corrected for multiple comparison, while (#) represents interaction effects between groups. The error bars represent 95% confidence interval. CW= cisgender women, TM= transgender men, CM= cisgender men, TW= transgender women.

**Table 1:**
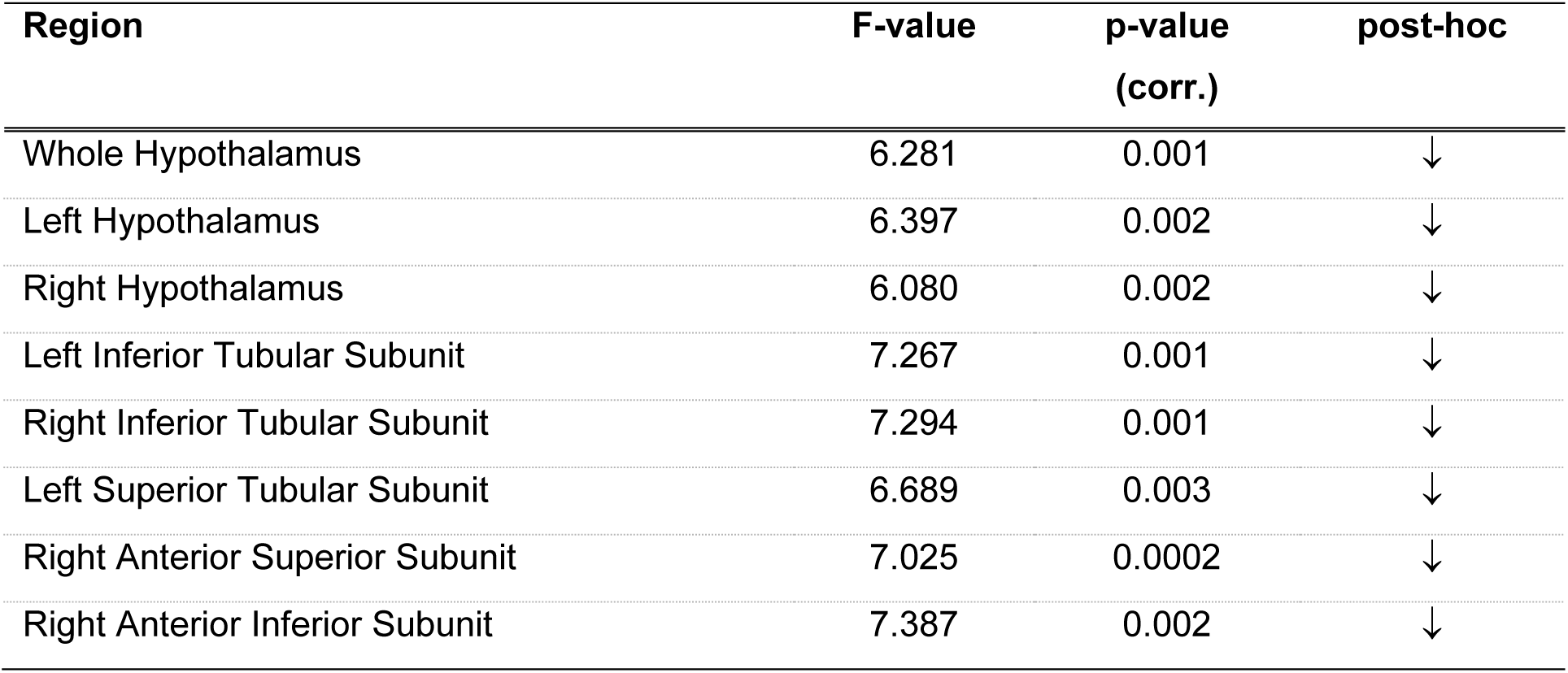
Significant results of hypothalamic volume and associated subfields in TW. F-values and corrected p-values of significant decreases in each region in transgender women (TW) after an interval of at least four months of estrogen and anti-androgen treatment are presented. (↓) represents post hoc decreases.

## 5. Discussion

In this study, the potential effects of estrogen and anti-androgen treatment on total hypothalamic volume as well as associated subfields were examined before compared to after an interval of at least four months of GHT in transgender individuals. Analyses were performed across four groups comprising TW and TM receiving GHT as well as untreated healthy controls (CW, CM). Group x time interaction effects were elucidated using repeated measures ANCOVA models. Here, we found significant reductions of hypothalamic volumes in TW receiving estrogen and anti-androgen treatment. Effects in the total hypothalamus have been attributed to the anterior and tubular subunits of the hypothalamus – more precisely, to the inferior tubular subunit bilaterally, the left superior tubular subunit, the right anterior inferior subunit, as well as the right anterior superior subunit.

Our result, showing a reduction in total hypothalamus volume in TW after at least four months of GHT are in line with an earlier finding of reduced total hypothalamic volume after four months of estrogen and anti-androgen treatment in TW (Pol et al., 2006). Though relevant, this study comprised a small sample size consisting of six TW and eight TM, presented uncorrected p-values and used an MRI segmentation method which was not directly comparable to the method used in our investigation. We were able to extend these findings by analyzing a larger sample size, presenting corrected p-values and using an automated MRI segmentation method, which aims to provide future comparability. Underlining our observations, findings showing volume reductions in multiple cortical and subcortical brain structures in TW after estrogen and anti-androgen treatment have been repeatedly published (Seiger et al., 2016; Zubiaurre-Elorza et al., 2014). In our study CW showed smaller hypothalamus volumes at the second MRI measurement compared to the first measurement. Although these results were not significant, it is interesting to note that *Pol et al*. made the same observations in their study (Pol et al., 2006). Estrogen and progesterone concentrations fluctuate during different phases of the menstrual cycle (Farage et al., 2009). These naturally occurring variations of hormone concentrations in the human body could affect brain morphometry as well. In fact, an exploratory analysis of naturally cycling women revealed volumetric differences of the total hypothalamus during different menstrual cycle phases (Chen et al., 2021). Also other studies showed variations of volumetric findings of the human brain in different menstrual cycle phases of naturally cycling women (De Bondt et al., 2013; Protopopescu et al., 2008). The cisgender women examined in our study could therefore have been in different phases of their natural menstrual cycle, which could reflect the volumetric differences found at the two timepoints and should be subject of future investigations.

A study exploring the effects of OCPs on human brain structure, reported smaller hypothalamic volume in healthy CW with OCP intake compared to naturally cycling CW (Chen et al., 2021). OCP mostly consists of estrogens and progestogens (Dragoman, 2014). Hence, the continuous intake of exogenously supplied estrogens in OCP users could have demonstrated a similarly decreasing effect on hypothalamic volume as discovered in the present study. Nevertheless, OCP use and estrogen treatment in GHT may not be directly comparable. Moreover, the authors provided no exact information on which kind of OCP was used. Therefore, drawing a comparison remains challenging. When focusing on other hypothalamic measures a study investigated effects of combined OCPs, consisting of estrogens and progestogens (Dragoman, 2014), in CW. Here, in the pill-free-phase transient microstructural and metabolic changes were observed in the hypothalamus (Baroncini et al., 2010). This phase is accompanied by a cessation of exogenously supplied estrogens and therefore a withdrawal of negative feedback mechanisms on the hypothalamo-pituitary-gonadal axis. Thus, an influencing impact due to differences in sex steroid hormone concentrations on hypothalamic measures could be assumed. The authors concluded that the elucidated changes might involve neuronal or glial remodeling of the hypothalamus in connection with gonadal axis activation (Baroncini et al., 2010), which in turn could take part in the explanation of our findings.

Studies investigating the impact of sex steroid hormones on brain structure revealed effects which occur in the developing brain and remain permanent, as well as influences in adulthood that are viewed as reversible. These are referred to as “organizational” or “activational” effects (Arnold, 2009; Gillies & McArthur, 2010). Activational effects on the hypothalamus have been elucidated in animal models. For example, in an ovine MRI study, baseline hypothalamic volume differences in males and females, which were described as sexually dimorphic, diminished after one month of gonadectomy (Barrière et al., 2019). Moreover, in earlier studies gonadectomized male mice which received estrogen and progesterone treatment showed significant reductions of volumes in hypothalamic nuclei in the medial preoptic area (Bloch & Gorski, 1988), as well as in the supraoptic area and the arcuate nucleus (Bloch & Gorski, 1988; Brawer et al., 1980). In “hormonally healthy” male mice estrogen and progesterone treatment did not lead to decreases in hypothalamic nuclei, therefore the authors deduced a protecting impact of accompanying high androgen levels on hypothalamic substructure (Bloch & Gorski, 1988). Taken together these findings might indicate that the effects in our study may firstly be referred to as activational effects and secondly, these effects might not only be present due to high estrogen levels but also because of simultaneously low testosterone levels. This hypothesis would be plausible considering an earlier described theory of a general anabolic and anticatabolic effect of testosterone on brain structure (Kranz et al., 2020; Zubiaurre-Elorza et al., 2014).

Another interesting aspect is how these volumetric findings could be functionally relevant in our study sample. As mentioned before the hypothalamus is known to be tightly integrated in a range of vital human functions which include endocrine and autonomic functions, as well as behavioral and emotional aspects. When considering the findings of our study, mainly the anterior and tubular regions of the hypothalamus were affected. This includes hypothalamic nuclei which are integrated in e.g., stress, arousal, sexual behavior, aggression, and sleep (Saper & Lowell, 2014). Interestingly, these hypothalamic nuclei were largely found to be influenced by estrogens in human and animal studies. To name a few, the ventromedial nucleus and PVN were attributed to be involved in the regulation of arousal and affective responses and have been found to be affected by estrogens during the menstrual cycle of CWs in an fMRI study (Goldstein et al., 2010; Goldstein et al., 2005). Moreover, the ventromedial nucleus is involved in aggressive behavior and was detected to play a substantial part in male and female aggression by expression of the estrogen receptor-*α* in mice (Hashikawa et al., 2017; Lee et al., 2014). Furthermore, estrogen has been found to influence dendritic spine density, a measure of synaptic connectivity, of the ventromedial hypothalamus in projection areas to the periaqueductal gray, which is crucially involved in female sexual behavior in a rat study (Calizo & Flanagan-Cato, 2002; Flanagan-Cato et al., 2001). Viewed from another perspective, transgender individuals revealed behavioral and hormonal changes during the course of GHT that may be related to functions of the mentioned nuclei. In that context, three months of estrogen and anti-androgen treatment was found to increase adrenocorticotropic hormone (ACTH) and cortisol levels after corticotropin-releasing hormone (CRH) administration in TW, which are values involved in stress response (Fuss et al., 2019). Moreover, sexual desire decreased during the first three months of GHT in TW, whereas it changed to be higher than baseline after 36 months of GHT (Defreyne et al., 2020). However, a link between aggression and GHT in transgender individuals remains inconclusive (Defreyne et al., 2019; Defreyne et al., 2018; Kristensen et al., 2021). Direct relationships between these relevant but exemplary presented structural and functional changes are difficult to pinpoint. Further studies investigating functional connections of hypothalamic subnuclei in transgender individuals would contribute to deepen the knowledge on the influence of sex steroid hormones on the human brain and potentially provide important information on neuronal changes during GHT to transgender individuals.

Some limitations of this study should be considered. The hypothalamus is a small structure in the human brain which exhibits low image contrast in MRI. Therefore, volumetric data in general remains scarce. With advances in this field however, improved analysis was performed and automated tools for MRI segmentation were presented, providing reproducibility and better comparability (Billot et al., 2020). In our study, we used the first automated tool, which performs segmentation of the whole hypothalamus as well as its subfields in T1-weighted MRI brain scans accurately and robustly (Billot et al., 2020). The hypothalamus, moreover, is anatomically located next to the third ventricle (Swaab et al., 1993). Studies investigating cortical and subcortical structures in transgender individuals found increased ventricle volumes in TW after four months of estrogen and anti-androgen treatment (Pol et al., 2006; Seiger et al., 2016; Zubiaurre-Elorza et al., 2014). These changes are discussed to be caused by surrounding grey or white matter structure variances during GHT (Seiger et al., 2016), which we cannot exclude being present in our findings. Nevertheless, we have tried to account for this effect by including TIV into our analysis. In addition, we examined data from two different studies conducted by our group, therefore our study sample was measured on two different 3T MRI devices. We corrected for multi-scanner imaging data using LongCombat (Beer et al., 2020), but cannot exclude influential factors. The effects seen in our study are most likely attributed to the estrogen and anti-androgen treatment TW received. However, we cannot exclude other confounding factors such as behavioral or mood changes as well as potential influences of other hormonal values. Nevertheless, we tried to rule out hormonal disruptors by adding relevant values into our statistic model.

## 6. Conclusion

Taken together, our results point toward a significant volumetric effect on the human hypothalamus and associated subfields after estrogen and anti-androgen treatment administered for an interval of at least four months in TW participants. Changes in hypothalamic volume have been mainly found in the anterior and tubular subfields of the hypothalamus – specifically, in the inferior tubular subunit bilaterally, the left superior tubular subunit, the right anterior inferior subunit, as well as the right anterior superior subunit. However, further studies investigating the effect of sex steroid hormones on human brain structure and functional connections of the hypothalamus and its subunits are needed to interpret these findings in a clinical sense.

## Data Availability

For reasons of data protection, the data is available on reasonable request from the corresponding author.

## 8. Author Contributions

The study was designed by RL and GSK. Medical support was provided by UK, PAH, MS, GMG, PBM and MEK. MRI measurements were performed by MBR, RS, BSD and MK. Administrative support was provided by VR. FreeSurfer analyses were performed by RS and BSD. Statistical analyses were conducted by MBR. MEK wrote the first draft of the manuscript. All authors contributed to the interpretation of the results, revised the manuscript and approved the final version.

## 10. Acknowledgments

We thank the graduated team members and the diploma students of the Neuroimaging Labs (NILs, headed by R. Lanzenberger), the members of the High Field MR Center, as well as the clinical colleagues from the Department of Psychiatry and Psychotherapy of the Medical University of Vienna for clinical and/or administrative support, especially S. Kasper, K. Papageorgiou, G. Gryglewski, T. Vanicek, M. Hienert, J. Unterholzner, A. Komorowski. Also, we would like to express our special thanks to the participants of the study, as it was their willingness to participate that made this study possible.

